# Emergency Response for Evaluating SARS-CoV-2 Immune Status, Seroprevalence and Convalescent Plasma in Argentina

**DOI:** 10.1101/2020.10.21.20216960

**Authors:** Diego S. Ojeda, María Mora Gonzalez Lopez Ledesma, Horacio Pallares, Guadalupe S. Costa Navarro, Lautaro Sanchez, Beatriz Perazzi, Sergio M. Villordo, Diego E. Alvarez, BioBanco Working Group, Marcela Echavarria, Kasopefoluwa Y. Oguntuyo, Christian S. Stevens, Benhur Lee, Jorge Carradori, Julio J. Caramelo, Marcelo J. Yanovsky, Andrea V. Gamarnik

## Abstract

We report the emergency development and application of a robust serologic test to evaluate acute and convalescent antibody responses to SARS-CoV-2 in Argentina. The assays, COVIDAR IgG and IgM, which were produced and provided for free to health authorities, private and public health institutions and nursing homes, use a combination of a trimer stabilized spike protein and the receptor binding domain (RBD) in a single enzyme-linked immunosorbent assay (ELISA) plate. Over half million tests have already been distributed to detect and quantify antibodies for multiple purposes, including assessment of immune responses in hospitalized patients and large seroprevalence studies in neighborhoods, slums and health care workers, which resulted in a powerful tool for asymptomatic detection and policy making in the country. Analysis of antibody levels and longitudinal studies of symptomatic and asymptomatic SARS-CoV-2 infections in over one thousand patient samples provided insightful information about IgM and IgG seroconversion time and kinetics, and IgM waning profiles. At least 35% of patients showed seroconversion within 7 days, and 95% within 45 days of symptoms onset, with simultaneous or close sequential IgM and IgG detection. Longitudinal studies of asymptomatic cases showed a wide range of antibody responses with median levels below those observed in symptomatic patients. Regarding convalescent plasma applications, a protocol was standardized for the assessment of end point IgG antibody titers with COVIDAR with more than 500 plasma donors. The protocol showed a positive correlation with neutralizing antibody titers, and was used to assess antibody titers for clinical trials and therapies across the country. Here, we demonstrate the importance of providing a robust and specific serologic assay for generating new information about antibody kinetics in infected individuals and mitigation policies to cope with pandemic needs.

**AUTHOR SUMMARY:** The development of robust and specific serologic assays to detect antibodies to SARS-CoV-2 is essential to understand the pandemic evolution and to stablish mitigation strategies. Here, we report the emergency development, production and application of a versatile ELISA test for detecting antibodies against the whole spike protein and its receptor binding domain. Over half million tests have been freely distributed in public and private health institutions of Argentina for evaluating immune responses, convalescent plasma programs and for large seroprevalence studies in neighborhoods and health care workers. We are still learning how and when to use serologic testing in different epidemiological settings. This program allowed us to produce large amount of high quality data on antibody levels in symptomatic and asymptomatic SARS-CoV-2 infections and generate relevant information about IgM and IgG seroconversion time and kinetics. We also present standardized protocols for antibody quantification as guidance for convalescent donor plasma selection in hospitals throughout the country for compassionate use and clinical trials. Here, we provide a framework for generating widely available tools, protocols and information of antibody responses for pandemic management.

## INTRODUCTION

The Americas have been profoundly impacted and have become the epicenter of Coronavirus Disease 2019 (COVID-19), as in October 13th more than 18 million infections and more than 550,000 deaths have been reported in the continent. Surveillance and testing are fundamental in controlling viral spread and understanding pandemic evolution. Detection of viral RNA by qPCR is the gold standard for diagnosis of acute infections. Measurement of humoral responses has been used as a complement to nucleic acid testing, for diagnosis of suspected cases with negative qPCR results, and for detection of acute or past infections in asymptomatic cases [1]. However, serology became an essential tool for the management of the pandemic, including seroprevalence assessment of immunity in the population, measurement of neutralizing antibody titers in convalescent patients and antibody response upon vaccination [2,3].

Antibody mediated immunity is thought to protect individuals from SARS-CoV-2 infection by interfering with viral entry and/or viral replication. Antibody responses appear within the first week of symptoms onset in about 30 to 40 % of infections and, in most cases, simultaneous or close seroconversion for IgM and IgG were observed [1,4]. Antibodies have been detected in more than 90% of infections after the third week of symptoms onset [5]. However, the duration and degree to which recovery from COVID-19 disease, or asymptomatic infection, confers prolonged immunity from reinfection is still unclear, even among individuals with high antibody titers [6,7]. We are still learning about the dynamic nature of antibody response linked to severe, mild, and asymptomatic COVID-19 manifestations and, as the pandemic progresses, algorithms and strategies to implement serologic testing in different epidemiological settings are under evaluation. For understanding this complex process, it is essential to have highly specific and sensitive serologic assays [8,9]. Based on the urgent need to attain reliable tests that measure antibodies to SARS-CoV-2, we redirected resources of a basic molecular virology laboratory, as part of a national task force for the emergency, for the development and production of an affordable and robust serologic assay for Argentina and neighboring countries. COVIDAR serologic test was generated early after the first COVID-19 case in Argentina, and over half million tests have been already produced and distributed for free in the country.

An important application of serology measurement is the assessment of humoral responses to vaccines and identification of plasma from convalescent donors for possible therapies [10–12]. During the initial stages of the COVID-19 epidemic in China, convalescent plasma therapy was used compassionately and has since been implemented in the United States and many other countries [13–17]. Randomized clinical trials to evaluate the usefulness of convalescent plasma in different stages of the disease are ongoing (https://clinicaltrials.gov). Success of this intervention likely increases with the antibody titer of the donor as recently shown [15,16]. Thus, it is important to screen potential convalescent donors to select plasma with the highest antibody titers. This screening can be accomplished by measuring the amount of antibodies by titration and defining the virus-neutralizing activity of the plasma. However, the lack of standardization and correlation between antibody levels and neutralizing capacity of antibodies complicates implementation and evaluation of plasma therapy protocols for COVID-19. Studies using SARS-CoV-2 in plaque reduction assays or pseudotyped particle-based systems indicate that plasma derived from convalescent patients has potent neutralizing activity related to IgG molecules recognizing the spike protein, suggesting that IgG antibodies against spike have high potential to fulfill neutralizing functions in vivo [18–21]. We developed standardized protocols for antibody quantification using COVIDAR IgG and provided data showing a positive correlation with neutralizing antibody measurements. This information allowed the implementation of protocols in hospitals throughout the country assessing plasma donors for compassionate use (https://www.groupcpc-19.com/) and for randomized multicenter clinical trials [22,23].

## RESULTS

Due to the urgent need for tools to cope with the pandemic, the Argentine Ministry of Science redirected scientific capacities to generate COVID-19 control means. In this context, the COVIDAR group developed a quantitative serologic test based on ELISA for SARS-CoV-2 infection, which gained emergency approval by the National Administration for Drugs, Food and Medical Devices (ANMAT) by May 4^th^. The assay uses two viral proteins, a trimer stabilized spike protein and the receptor binding domain (RBD), expressed in human cells. Combining these proteins in one assay provided higher sensitivity and reproducibility than using each protein in independent plates for IgG and IgM testing [24].

For this study, four different cohorts of patient samples were used. First, a full data set of 535 acute or convalescent COVID-19 qPCR positive serum samples; second a data base of antibody measurements and specific information of suspected COVID-19 with 3500 entries; third, 1074 serum samples from longitudinal studies of a cohort of 93 patients; and fourth, 561 COVID-19 convalescent plasma from potential donors.

IgG and IgM responses were initially evaluated in 535 qPCR positive patients (Figure 1A). Samples were grouped according to days of symptoms onset (SO). In the early phase, within 7 days of SO, the IgG sensitivity was 35% (Figure 1A). The sensitivity for IgG detection increased from 72% to 74% between 2 and 3 weeks, and seroconversion increased up to 90.4% after 3 weeks. In addition, within 100 samples with >45 days of SO, 5 did not show presence of IgG, indicating that 5% of the patients displayed undetectable levels of IgG or they were non-responders up to 45 days. Regarding IgM, within 7 days of SO, sensitivity was 46%. IgM levels were highest between 15 and 21 days, and although IgM positivity increased after 3 weeks, the antibody level observed by optic density (OD) at 450 nm clearly decreased (Figure 1A).

**Figure 1.**
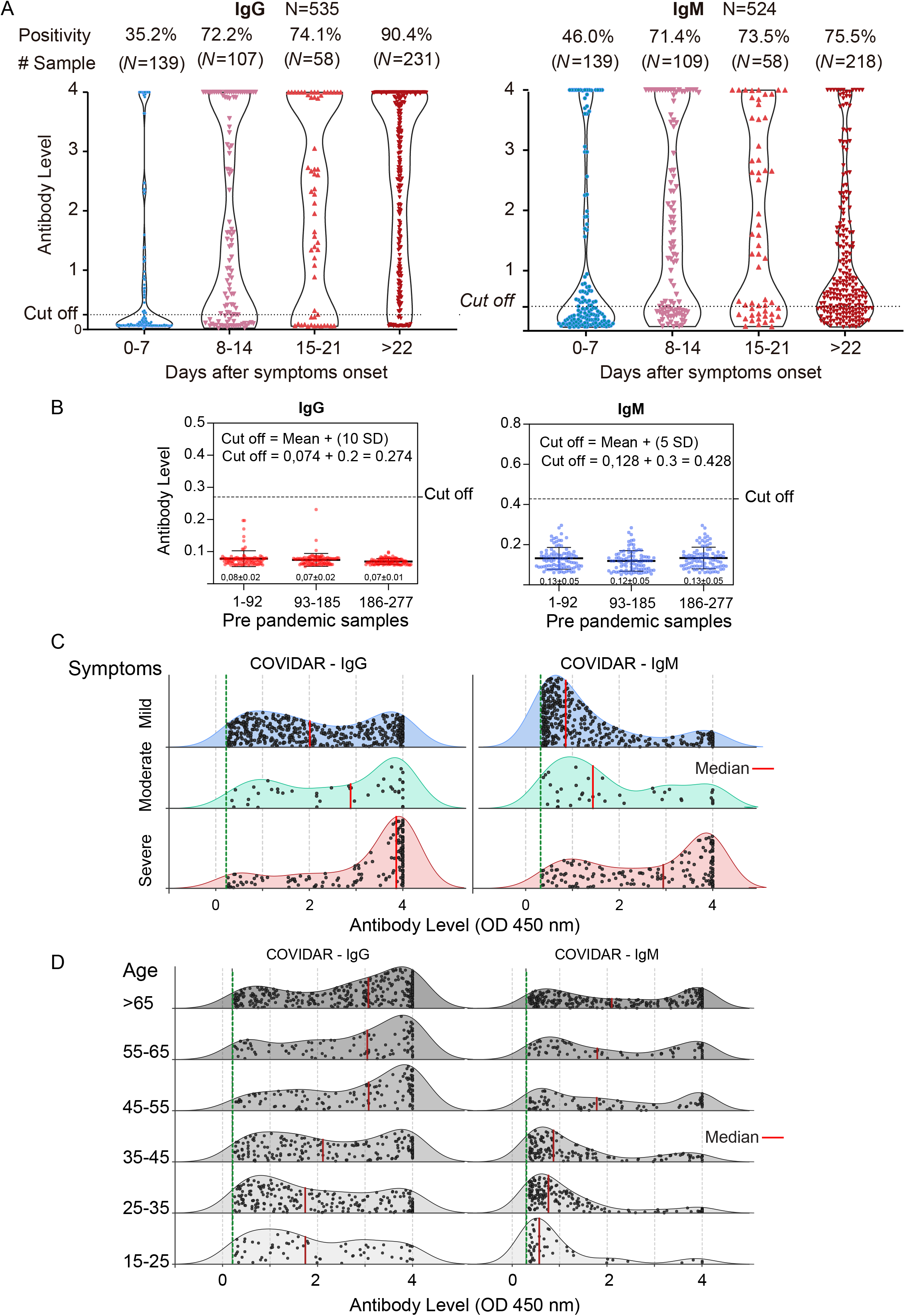
Antibody responses against SARS-CoV-2. **A**. Levels of specific IgG and IgM antibodies against SARS-CoV-2 versus days of symptoms onset in 535 patient serum samples. The positivity rate and sample size for each week are indicated on the top. The cut-off is also shown. **B**. Levels of IgG and IgM antibodies against SARS-CoV-2 in 277 pre-pandemic serum samples. The cut-off definition is shown. **C**. Comparison of the levels of IgG and IgM antibodies against SARS-CoV-2 from patients with severe, moderate and mild symptoms. For each plot, the red lines indicate median values. **D**. Comparison of the level of IgG and IgM antibodies against SARS-CoV-2 from patients of different ages. The number of samples analyzed for C and D was 1379 and 976, respectively.

Assessment of 277 pre-pandemic samples, including acute infections of other pathogens (dengue, HCV, HIV and seasonal coronavirus), were performed. The cut off for IgG and IgM were defined to maximize specificity, ensuring no false positives. Thus, the cut off was defined as the mean of negative control plus 10 or 5 standard deviations for IgG and IgM, respectively (Figure 1B). High specificity of COVIDAR IgG has been supported by baseline seroprevalence studies of heath care professionals (HCP). In this regard, seroprevalence in HCP at region VIII of Buenos Aires Province, performed in June 2020, showed 0.74% of IgG positivity [25].

A data set of IgG and IgM, including 1379 samples of acute and convalescent infections with complete medical records, was used to analyze distribution of age, gender and symptoms. No significant differences concerning gender were observed. The amount of patients with high IgG levels in the group of severe symptoms was larger than those in the moderate and mild groups. The distribution of IgG levels for patients with severe symptoms were significantly different from those with mild symptoms (Wilcoxon rank sum test p-value<0.0001). The median OD level at 450 nm for IgG was 2, 2.9 and 3.9 for mild, moderate and severe groups, respectively (scale up to 4). This trend of antibody levels with symptoms was also evident for IgM, median 0.8, 1.4 and 2.9 for mild, moderate and severe groups, respectively (Figure 1C). The distribution of IgM levels for patients with severe vs mild symptoms were significantly different (Wilcoxon rank sum test p-value<0.0001). Antibody measurements also correlated with age, covering a range from 16 to more than 65 years showing the highest antibody levels associated with older COVID-19 patients (Figure 1D). Antibody level distribution significantly differed between patients that were < than 45 versus those that were > than 45 years old (Wilcoxon rank sum test p-value<0.0001).

Serological courses were also followed in a separate cohort of 93 hospitalized patients, most of which (90 patients) underwent seroconversion during the follow-up period (Figure 2A). From this, 1074 antibody determinations were performed. We represented the seroconversion time for IgM and IgG for each patient, as the first moment in which antibodies were detected. In most cases, synchronous seroconversion of IgG and IgM was observed (67 patients, 72%). IgM seroconversion was earlier than IgG in 24% of the patients, while IgG appeared earlier than IgM in 9% of the patients (Figure 2A). This longitudinal study indicates that in the first week of SO 34% of the patients were IgG positive and 38% IgM positive, in agreement with data provided in Figure 1, obtained with a different data set and analysis. In addition, considering patients that were followed for at least 45 days of SO, the seroconversion rate in this cohort was 96.7%.

**Figure 2.**
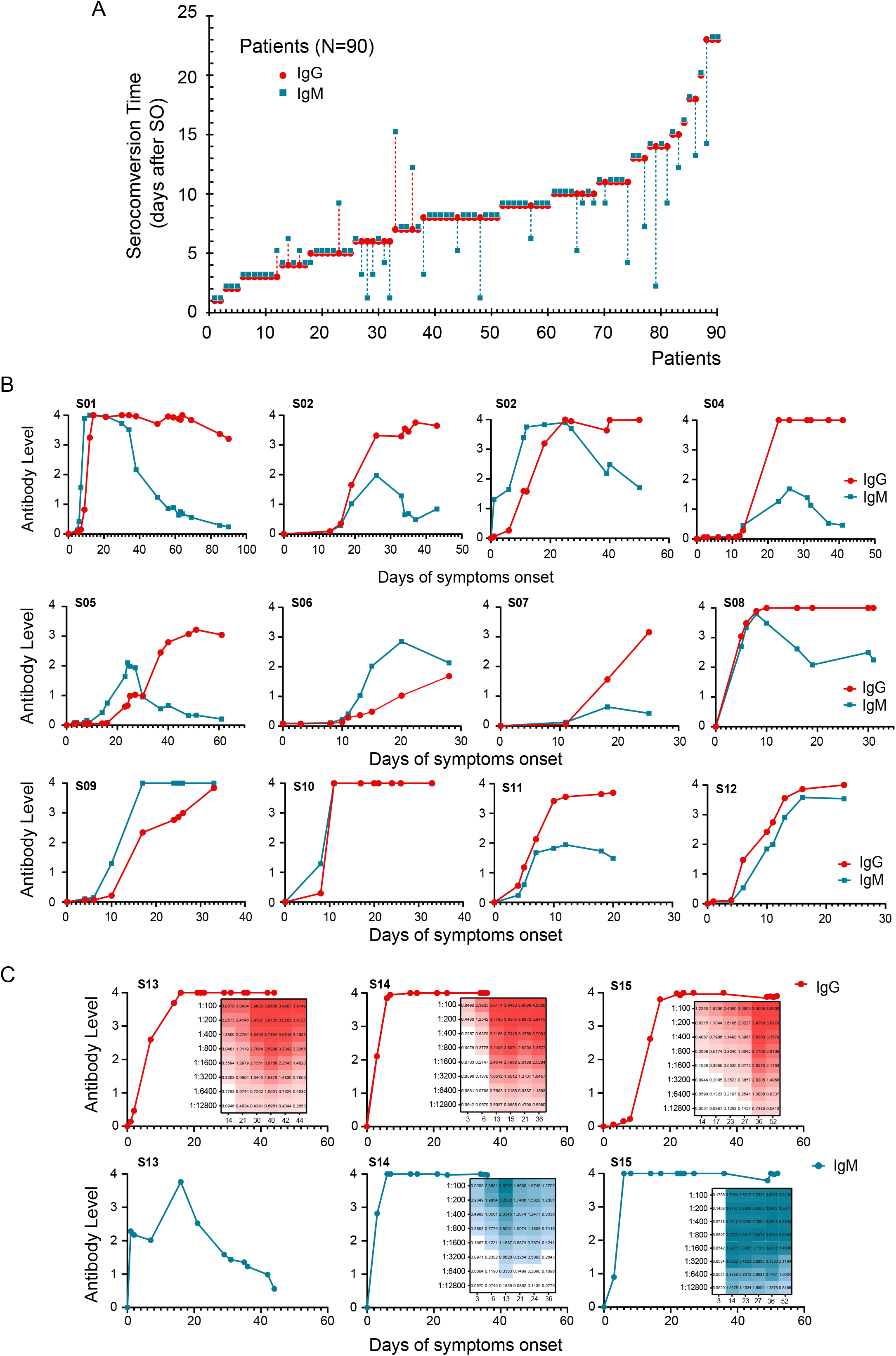
Longitudinal antibody measurements of SARS-CoV-2 infected patients. **A**. Antibody responses of 90 patients, initially seronegative, were followed as a function of time until seroconversion. The plot indicates the time at which antibodies appear for the first time for each patient. IgM and IgG seroconversion time is shown in different colors (blue and red, respectively). Three types of seroconversions were observed: synchronous (IgG and IgM together), or consecutive (either IgM or IgG first). The time separation between the two seroconversions are indicated by a dash line. **B**. Representative longitudinal IgG and IgM antibody measurements of symptomatic SARS-CoV-2 infected patients show large humoral response heterogeneity. Each plot represents a single patient followed as a function of time of symptoms onset. **C**. Representative examples of longitudinal antibody measurements with different IgM profiles of patients with severe symptoms. Insets indicate heats maps of antibody titrations (from 1/100 to 1/12800).

For other infectious diseases, information regarding duration of IgM is relevant for identification of possible active infections. In our cohort of COVID 19 subjects, longitudinal studies of antibody responses showed a considerable heterogeneity of IgM kinetics (Figure 2B). Analysis of IgM profiles of the data set shown in Figure 2A indicate that in 60% of the cases IgM levels declined before 30 days of SO, while in 40% of the patients it remained at high levels after 30 days, highlighting a wide range of IgM responses and kinetics in different patients (Figure 2B). Specific examples of longitudinal measurements of patients with severe symptoms indicate that while IgG levels plateau at high levels and remained, in most cases, at elevated titers between 40 and 60 days of SO, IgM dynamics were also very diverse (Figure 2C). IgM titrations indicate that, while in some patients the antibody levels abruptly or slowly dropped before 30 days, in other patients IgM levels remained high (titers above 1/12,000), even at 60 days of SO (Figure 2C), suggesting that IgM detection should be used with caution in SARS-CoV-2 infections.

Serology testing has been performed systematically in HCP and nursing homes using COVIDAR in asymptomatic populations for surveillance. Data indicate that asymptomatic infections were detected by IgG and IgM testing, though at lower levels compared with symptomatic patients (Figure 3A). In addition, longitudinal studies of SARS-CoV-2 infections in the absence of symptoms also indicated simultaneous or close seroconversion of IgM and IgG (Figure 3B).

**Figure 3.**
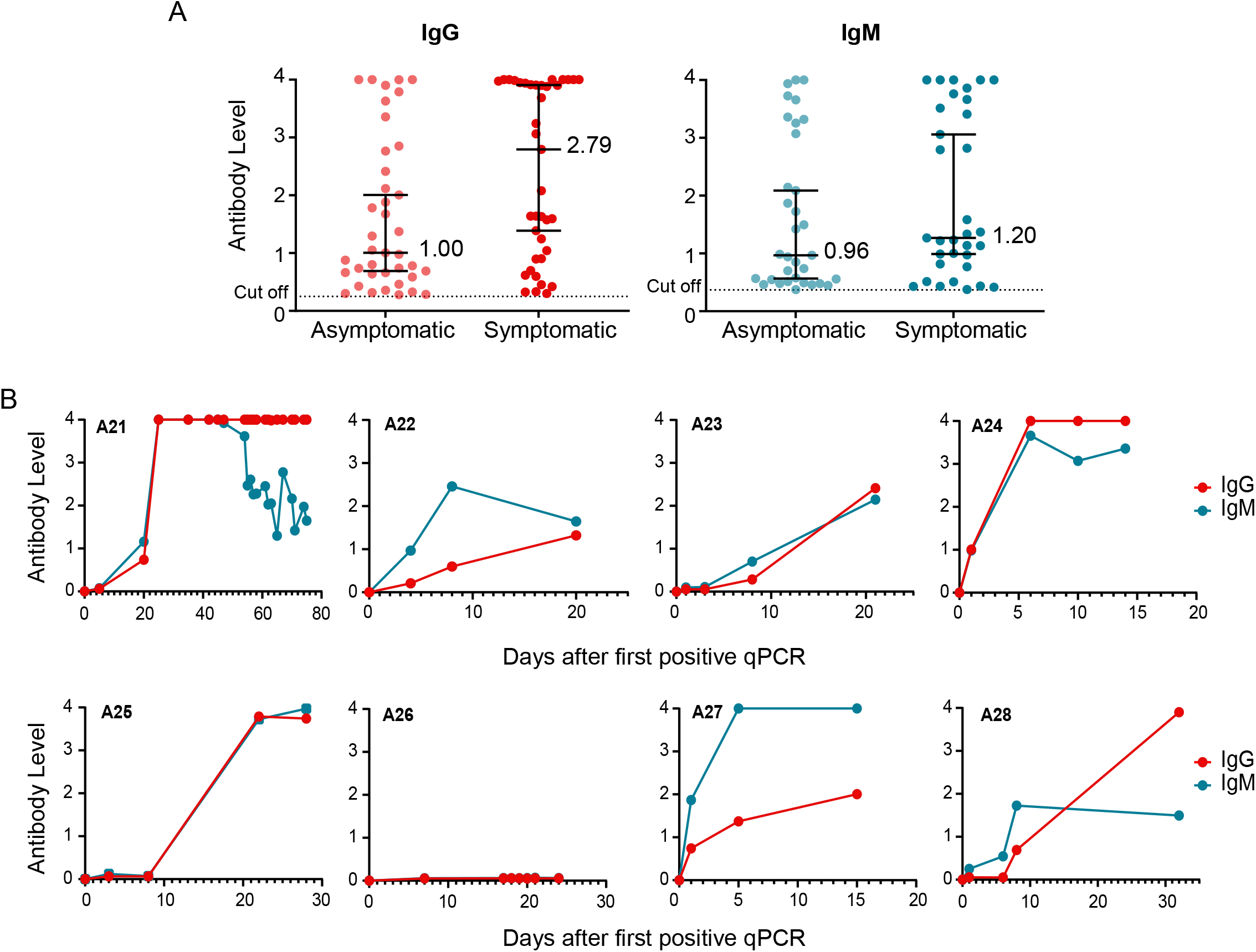
Antibody responses of asymptomatic SARS-CoV-2 infected patients. **A**. Comparison of virus-specific IgG and IgM antibody levels in asymptomatic (*n* = 40) and symptomatic patients (*n* = 40), during acute SARS-CoV-2 infection, are shown. The median is indicated in each case. **B**. Representative examples of longitudinal antibody measurements of asymptomatic SARS-CoV-2 infected patients. IgM and IgG levels are represented as a function of time of the first qPCR positive test for each patient.

**Figure 4.**
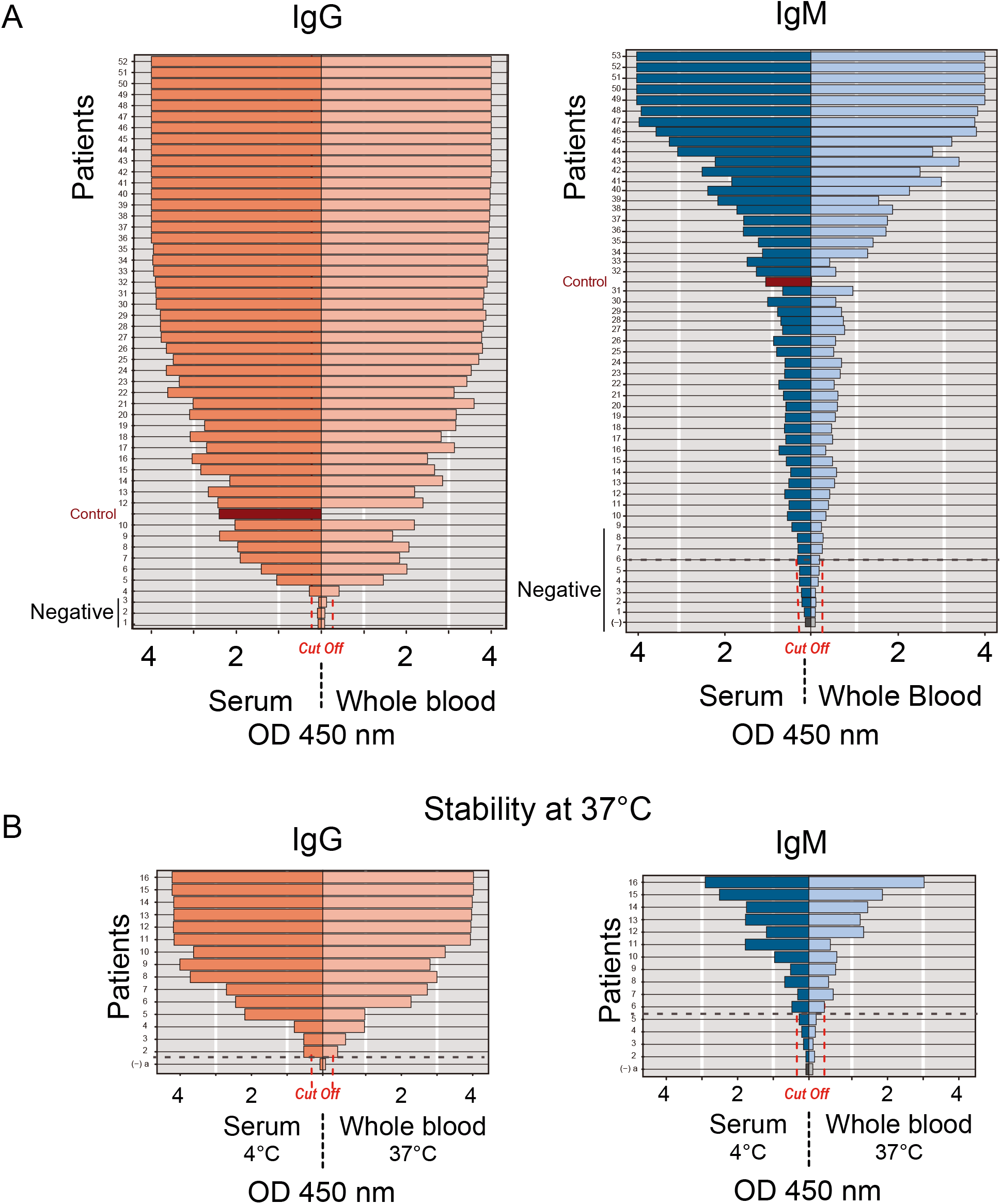
Validation of finger prick sampling for COVIDAR ELISA test. **A**. Antibody measurements of paired serum and whole blood samples from the same patient were performed using COVIDAR IgG and IgM. For each case the positive control (red) and cut off are indicated. **B**. Stability of whole blood sample at 37°C in Serokit for field SARS-CoV-2 serosurvey studies. Antibody measurements of paired whole blood and serum samples from 19 patients that were kept for one week at 37°C in Serokit or 4°C, respectively.

### Whole blood antibody testing by ELISA assay validated for large scale seroprevalence studies

Well-designed serosurveys are essential to determine how prevalent COVID-19 is in the general population, in selected subsections of the population (such as health care workers), or in specific risk groups. Highly specific and sensitive assays are desirable for accurate seroprevalence studies; however, it is also necessary to have methods that are suitable for simple and fast sample collection. ELISA assays use venipuncture, and require refrigerated storage of samples during transportation, making the assay incompatible with massive testing. To overcome this limitation, we adapted the COVIDAR IgG and IgM assays to work with whole blood samples obtained by finger prick. A self-contained Serokit was validated for storage and transportation of whole blood using a glycerin based preservation system [26]. Similar methods have been extensively used in serosurvey studies of Chagas disease conducted in rural areas of Argentina [27].

To validate the process, 68 paired serum and whole blood samples from confirmed SARS-CoV-2 qPCR positive patients were used. All serum specimens were kept at 4°C, while whole blood samples collected from the same individuals were divided as follows. Fifty two whole blood samples were kept at ambient temperature and, to evaluate stability, 16 samples were placed at 37°C for 7 days using the Serokit preservation system. IgG and IgM antibody reactivity from serum samples showed 100% agreement with those from the same individual’s whole blood samples stored using Serokit, either at room temperature or at 37°C (Figure 3).

The validation of finger prick capillary whole blood sample application with Serokit storage widened COVIDAR use to field serosurveys. Until now, seroprevalence studies in nine slums of Buenos Aires, and surveillance studies among HCP in 30 hospitals have been conducted [28–30].

### Standardized protocols for IgG quantification that correlate with neutralizing activity

An important limitation to evaluate the quality and quantity of antibodies present in COVID-19 convalescent plasma is the lack of tests that can be easily performed in each institution for possible therapies and clinical studies. Previous analysis using serologic test assessing IgG titers directed to RBD and/or spike have shown a positive correlation with antibody neutralization [7]. Due to the urgent need for tools, we validated an IgG titration protocol using COVIDAR and performed correlations with neutralizing antibody titers. Protocols and COVIDAR kits for IgG titration were freely distributed in private and public institutions in 20 out of 23 provinces of the country, in which more than 100,000 tests have been already performed.

Five hundred and sixty-one convalescent donor plasma samples were tittered in our laboratory for standardization. A distribution analysis of the amount of donors and IgG levels indicate that more than 80% of the samples displayed spike specific end point IgG titers above 1/400. Hyper-immune plasmas with titers as high as 1/400,000 were observed (Figure 5A). A significant difference of IgG titers was observed between donors that experienced mild symptoms and those that experienced moderate or severe symptoms, median of 1/800 and 1/6400, respectively (p=0.0001) (Figure 5A). Full titration curves are shown in heat maps to illustrate this distribution (Figure 5B).

**Figure 5.**
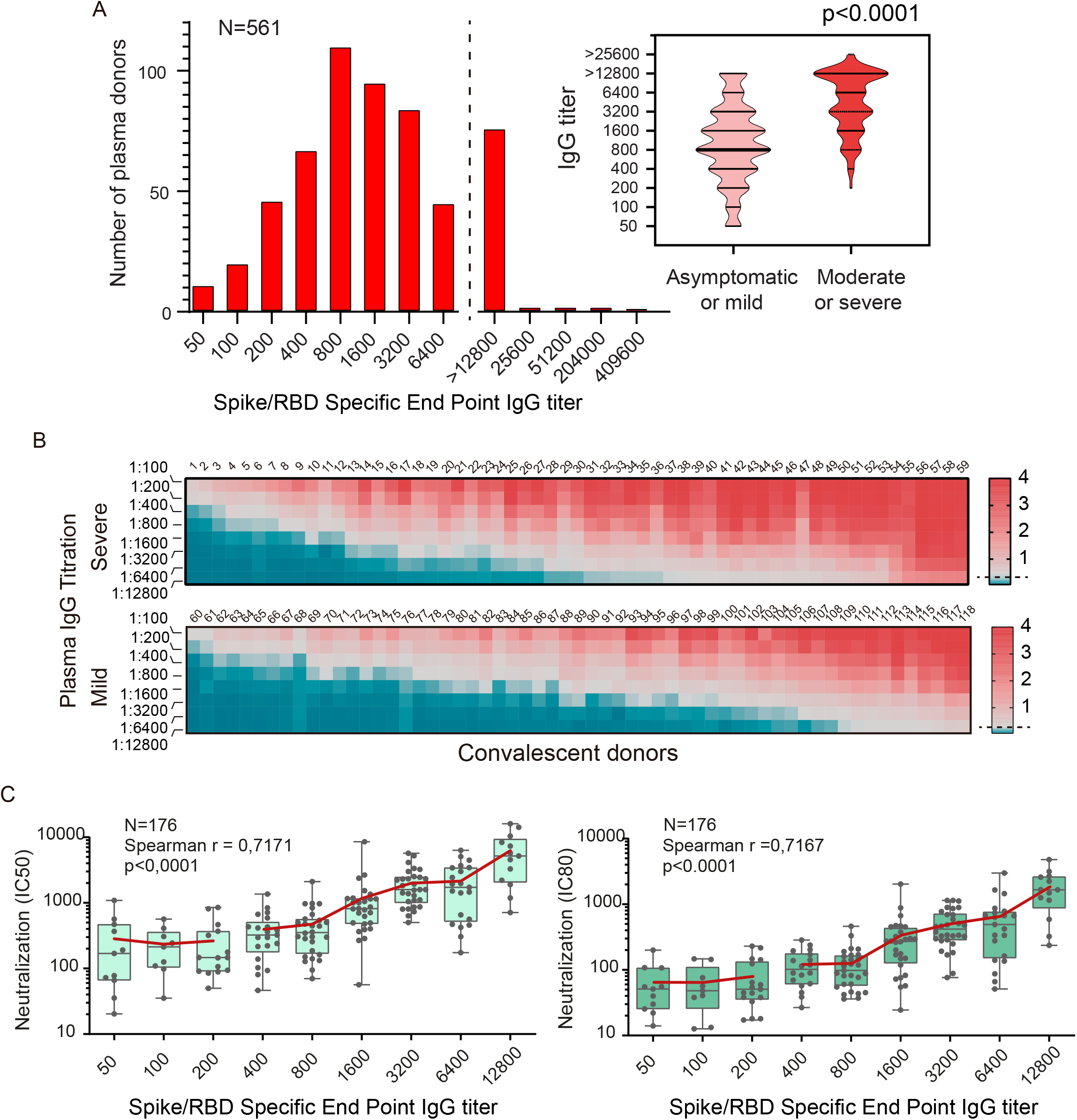
Quantification of IgG levels in COVID-19 convalescent patients and correlation with neutralizing activity. **A**. IgG titers were defined by end dilution with COVIDAR ELISA test in convalescent-phase COVID-19 patients who were discharged from the hospital (n = 561). On the right, plot of titer distribution showing significant differences (p <0.0001, Mann-Whitney test) between samples from donors that have experience mild or severe symptoms. **B**. Full IgG titrations curves of plasma sample from donors that recovered from severe or mild COVID19 were used to generate heat maps. Red represents high and blue low antibody levels as indicated on the right. Samples were sorted by increasing titers. **C**. Correlation of IgG end point titration and neutralizing antibody titers in COVID-19 patients measured by pseudovirus CoV2pp (n = 176). The reciprocal of inhibition concentration 50% and 80%, IC50 and IC80 respectively, are correlated to the reciprocal end point IgG titers. In the inset the r Spearman and p values from linear regression are shown. Boxes indicate the median, and the red line the mean of IC50 or IC80 for each IgG titer.

The data using the IgG titration protocol was correlated with virus-neutralizing activity using SARS-CoV-2 pseudotyped VSV particles (CoV2pp) [31]. Plasma from 176 potential donors were used to assess neutralizing activity. Neutralization curves were fit using a 4-parameter logistic regression model and used to determine 50 and 80% CoV2pp absolute infection inhibition concentration (AbsIC50 and AbsIC80, respectively) by measuring luciferase reporter activity encoded in the CoV2pp. AbsIC50 and AbsIC80 values were correlated with spike specific end point IgG titers (r=0.717, p<0.0001 and r=0.716, p<0.0001 respectively, Figure 5C).

Convalescent plasma samples with low IgG titers between 1/50 to 1/200, displayed detectable neutralizing antibodies, with IC80 from the reciprocal of 16 to 132, highlighting that all COVID-19 convalescent individuals with positive anti-spike IgG showed neutralizing antibodies (Figure 5C). A positive correlation between spike binding IgG titers and CoV2pp infection neutralizing activity was observed with convalescent donor plasma titers above 1/200 (Figure 5C). Plasmas with high spike IgG titers invariably displayed high neutralization activity estimated by IC50 or IC80.

We conclude that using end point titrations for total IgG against spike with COVIDAR is suitable for large scale studies, especially in areas of limited resources, as surrogate of neutralization potency and guidance for convalescent donor plasma selection.

## DISCUSSION

The development and appropriate application of serologic assays to detect antibodies to SARS-CoV-2 are essential to determine the pandemic evolution and stablish mitigation policies. Here, we developed and produced a widely available serologic tool to assist health authorities for pandemic management. Protocols and reagents were distributed to evaluate immune status of hospitalized patients, surveillance of at risk groups, seroprevalence studies in different populations and evaluation of plasma from convalescent donors for possible therapies. The biggest challenge still stands regarding how to deploy this tool in a strategic manner to bring communities out of the current suffering.

We provided a robust serologic test for health institutions throughout the country and generated highly needed information of humoral responses of acute and convalescent SARS-CoV-2 infections. Specific IgG and IgM responses were found to be highly heterogeneous among different individuals. We observed early seroconversion within the first week of SO in at least 34% of the patients (Figure 1 and 2). This raised the question of whether IgM and/or IgG seroconversion time indicates the end of infectiousness or whether early antibody appearance may overlap with active infection and possible transmission. Although this is an issue that requires further attention, periodic serology testing has been useful for guiding focused qPCR testing, asymptomatic case identification and contact tracing.

The fraction of asymptomatic but infectious cases is a critical epidemiological characteristic that modulates pandemic potential. Viral load and antibody responses in asymptomatic individuals are both important to understand transmission and pandemic extension. It is still unclear how many people carry SARS-CoV-2 infection asymptomatically. It has been shown that the ratio symptomatic to asymptomatic cases changes in different settings. Ratios from 2:1 to near 1:8 symptomatic to asymptomatic cases have been reported [28,32,33]. The factors that influence this large variation are still unknown. One concern is that the low level of antibodies found in asymptomatic cases may lead to their underestimation [34]. Our studies show wide individual variations in the antibody response and kinetics in both symptomatic and asymptomatic cases. We found that asymptomatic individuals displayed lower overall antibody levels than those observed in COVID-19 symptomatic patients (Figure 3). A positive correlation between disease severity and levels of IgG and IgM in acute and convalescent stages was also observed (Fig 1C and 5A), in agreement with previous studies [6,35]. Another important issue is the duration of IgM antibodies. Our longitudinal study using 93 infected individuals showed that IgM levels wane in 60% of the cases within 30 days of SO (or first qPCR detection for asymptomatic cases). However, IgM levels were still detected after 30 or even 60 days in 40% of the cases, suggesting that IgM detection does not necessarily reflect a recent infection.

Another important question is whether all infected individuals mount a robust antibody response to SARS-CoV-2 infection. In this regard, we found antibodies in 90% of the qPCR positive cases after 3 weeks of SO and, considering patients that were followed for at least 45 days, the seroconversion rate reached 95%. Thus, at least 5% of infected individuals resolved the infection with undetectable levels of antibodies. This observation could be assigned to low antibody responses, below the detection limit, or to non-responder individuals. In this regard, previous studies have shown the relevant role of T cell responses in infection resolution [36,37].

Factors that affect the spread and dynamic of the virus in different geographic, demographic, and socioeconomic areas are still unclear. Reliable reagents to perform large and periodic serosurveys are urgently needed. Here, we provided a tool for using whole blood by finger pricking, followed by IgG and IgM tests, to facilitate large sampling together with a robust ELISA assay. This approach has been used in different neighborhoods and defined at-risk populations in Argentina with SARS-CoV-2 prevalence from less than 1 to more than 50% [28,29], supporting the convenience and utility of the application.

It is important to stress the relevance of widely available quantitative serologic assays. Passive antibody transfer is a treatment strategy that has been used for COVID-19, including plasma and purified immunoglobulins derived from COVID-19 convalescent donors. Although the efficacy of convalescent plasma treatment remains uncertain, recent reports indicate a clinical benefit associated with high-titer units administered early in the course of the infection [16]. In this regard, an important limitation has been to obtain harmonized antibody titers information used in different clinical trials and therapeutic applications. By tittering more than 500 convalescent plasma samples, we showed a diversity of titers, which correlated with donor disease severity (Figure 5A). We provided a standardized protocol, widely distributed and used in Argentina for quantitative IgG measurements. A positive correlation between IgG quantitation, using COVIDAR, with neutralizing activity measured by a pseudotyped virus was shown (Figure 5C). This protocol has been useful to normalize quantifications at hospitals and heath institutions using plasma for compassionate therapies and for different clinical trials [22,23]. Although IgG titers correlated with neutralizing activities, the relevant open question is how they correlate with protection [38,39].

While the pandemic still progresses, scientists, public health workers, and policy makers are being challenged to create new strategies based on evidence and experiences in different parts of the world. A consensus has emerged that serological testing provides an essential tool in the pandemic response, nevertheless, serology testing strategies should be dynamic and adaptable to specific needs and resources available in different regions. Our work provides widely and freely available robust serology reagents, protocols, and data on humoral responses of SARS-CoV-2 infected individuals that hopefully helps finding better control measures for the current pandemic.

## EXPERIMENTAL PROCEDURES

### SARS-CoV-2 protein expression

The ectodomain, soluble version of the SARS-CoV-2 spike protein (residues 1–1208 of GenBank: MN908947) including a T4 foldon trimerization domain, a GSAS substituted at the furin cleavage site (residues 682–685), and an octahistidine tag was cloned into a pCDNA mammalian expression vector. A SARS-CoV-2 version S-2P with substitutions at 986 and 987 was originally used. However, due to the difficulty of purifying the necessary amount of spike protein for producing the ELISA plates, substitutions that increased protein yields and stability were incorporated. In this regard, a variant with four additional proline substitutions was generated, HexaPro variant, as recently described [40]. The sequence of the receptor binding domain (RBD, amino acid 319 to 541, RVQP….CVNF) along with the signal peptide (amino acid 1-14, MFVF….TSGS) plus a hexahistidine tag was obtained from the Krammer laboratory [41] and subcloned into a pCDNA human expression vector.

For protein expression, FreeStyle293-F (293-F) cells or adherent HEK293-T (293-T) cells were maintained at 37°C with 8% CO_2_ and 5% CO_2_, respectively. 293-F cells were grown and transfected in Expi293™ expression medium and 293-T cells were grown in D-MEM high glucose supplemented with 10% Fetal Bovine Serum and Penicillin/Streptomycin 1% and transfected in Opti-MEM reduced serum media. Plasmids encoding Spike and RBD were transiently transfected using polyethyleneimine hydrochloride. Transfected 293-F suspension cultures were maintained at 32° and transfected 293-T cells were maintained at 37°C. The cultures supernatants were harvested at five days after transfection by centrifugation of the culture at 4000 g for 20 minutes at 4°C, new medium was added and a second harvest was performed after four days. For FreeStyle293-F cells viability was monitored and kept above 75% at all times.

### Protein purification

Culture supernatants were applied using a peristaltic pump to HisTrap excel columns (GE Healthcare) with a flow rate of 2-3 ml/min, using 1 ml of media for each 100 ml of supernatant. Columns were washed with 10 volumes of buffer A (NaH_2_PO_4_ 57,5mM, NaCl 300 mM, pH 7.8) and next with 60 volumes of buffer A with 10 mM imidazol. Then, columns were connected to an HPLC system (Kanuer) and washed at 2 ml/min with the last buffer until absorbance at 280 nm reached a value below 0.005. Proteins were eluted with a 12 minute linear gradient from 10 to 1 M imidazole in buffer A. Fractions of 1 ml were collected and analyzed by SDS-PAGE. Buffer of fractions containing protein was exchange to PBS with NAP-5 columns (GE Healthcare) and pooled. Typical yields were around 6 and 45 mg/l for spike and RBD, respectively, with a purity level of at least 98 %.

### Enzyme-linked immunosorbent assays development

The ELISA protocol was adapted from previously established protocols used for Chagas disease by Laboratorio Lemos S.R.L. [42], and for SARS-CoV2 by the Krammer Laboratory [41]. Ninety-six well high binding plates (Jet Biofil, Guangzhou, China) were coated overnight at 4°C with 75 ul per well of a solution containing 100 ng of a mixture of full-length trimeric spike and RBD proteins in carbonate buffer, pH 9.6. The next day the coating solution was removed by washing three times with phosphate-buffered saline containing 0.05% Tween (PBS-T), pH 7.4. Serum samples diluted in PBS-T containing 0.8% casein were added to the wells (100 μl of a 1:100 dilution for IgM and 200 μl of a 1:50 dilution for IgG determination), and incubated for 1 h at 37°C. For end point titrations, samples were serially diluted in IgG SARS-CoV-2 negative serum or fetal bovine serum (FBS), added to the plates and incubated for 1 h at 37°C. Following a washing step with PBS-T, 100 μl of diluted horseradish peroxidase (HRP)-conjugated with goat anti-human IgM (Sigma), or with mouse anti-human IgG antibodies (BD pharmingen), was added to the wells and incubated for 30 min. at 37°C. Subsequently, the plates were washed with PBS-T, and the peroxidase reaction was visualized by incubating the plates with 100 μl of TMB solution for 30 min at 37°C. The reaction was stopped by adding 100 μl of 1M sulfuric acid, and optical densities (OD) were immediately measured at 450 nm. For whole blood analysis, 50 μl of blood were preserved in collecting tubes containing 90 μl of buffered glycerin from the Serokit system [26]. Stored samples were then diluted in PBS-T containing 0.8% casein for quantification of IgG or IgM levels.

### Serum and plasma samples

Human plasma and serum samples were obtained from a number of different sources following the subsequent inclusion criteria: people aged over 16 years, of any sex who presented a diagnosis of infection or suspected of SARS COVID-19 infection in acute or convalescent phase. Confirmed or suspected cases met the criteria of The National Ministry of Health of Argentina (https://www.argentina.gob.ar/salud/coronavirus-COVID-19/definicion-de-caso). De-identified samples from the following health institutions were received: BioBanco de Enfermedades Infecciosas (BBEI), Centro de Educación Médica e Investigaciones Clínicas “Norberto Quirno” sede Saavedra (CEMIC), Hospital de Clínicas José de San Martín, Hospital de Infecciosas Fransisco Javier Muñiz, Hospital General de Agudos Dr. I. Pirovano, Hospital General de Agudos Parmenio Piñero, Hospital Italiano, Hospital Militar Central Cirujano Mayor Dr. Cosme Argerich, Hospital Naval Dr. Pedro Mallo, Hospital Universitario Austral, Hospital Sirio Libanes, Sanatorio de La Trinidad Mitre, Sanatorio Dr. Julio Méndez, Sanatorio Franchin, Sanatorio Güemes and Swiss Medical.

A pre-pandemic serum panel of samples selected based on the date of collection (July and September 2019) was provided by the Blood Bank of the Hospital de Clínicas José de San Martín. Positive and negative controls were donated from the CEMIC blood bank. Longitudinal studies were performed using samples from de-identified patients from Sanatorio de La Trinidad Mitre and Hospital de Clínicas José de San Martín (n=118). Samples were taken every 2 to 4 days until the patient was discharged from the hospital. Additional samples were collected when patient assisted for controls. Samples were collected between 1 to 90, or more days, of SO.

Asymptomatic patients identified by contact tracing of COVID-19 positive patients were obtained from BBEI COVID-19 collection, Hospital de Clínicas José de San Martín, Sanatorio de La Trinidad Mitre and Swiss Medical. Paired whole blood and serum samples used for Serokit validation were obtained from the BBEI COVID19 collection.

The study was conducted in accordance with the Declaration of Helsinki, and was reviewed and approved by the Bioethics Committee of Fundación Instituto Leloir (HHS IRB # 00007572) (CBFIL Protocol #35). All participants provided written informed consent prior to the study. As for participants under 18, informed consent was not obtained since samples were de-identified.

Sample collection and processing were performed from April 2020 to September 2020.

### Neutralization assay

Neutralization assays were carried out with SARS-CoV-2 pseudotyped particles (CoV2pp), generated in Benhur Lee’s laboratory [31]. CoV2pp carries vesicular stomatitis virus as viral backbone, bearing the Renilla luciferase gene in place of its G glycoprotein (VSVΔG-rLuc), and expresses SARS-CoV-2 Spike protein on its envelope.

ACE2 and TMPRSS2 expressing 293T cells (293T-ACE2+TMPRSS2 clone F8-2), were used for these assays [31]. Cells were maintained with DMEM high glucose with 10% FBS and were seeded in a 96-well plate the day before infection.

Patient sera were heat inactivated at 56°C for 30 minutes and serially diluted in DMEM high glucose with 10% FBS. Serum neutralizations were performed by first diluting the inactivated sample 8-folds and continuing with a 2-fold serial dilution. A pre-titrated amount of pseudotyped particles (diluted to give approximately 5×10^5^ Relative Luminescent Units) was incubated with a 2-fold serial dilution of patient sera for 30 minutes at room temperature prior to infection. Approximately 20 hours post-infection cells were processed for detection of luciferase activity. Cells were lysed and transferred to white, F-bottom 750 Lumitrac plates (Greiner, 655074). Plates were read via the GloMax® Navigator Microplate Luminometer (Promega, GM200) using the Nano-Glo® Luciferase Assay System (Promega, 752 E6110).

Absolute inhibitory concentrations (absIC) values were calculated for all patient sera samples by modeling a 4-parameter logistic (4PL) regression with GraphPad Prism 8. The 4PL model describes the sigmoid-shaped response pattern. For clarity, it is assumed that the response can be expressed so that the slope increases as the concentration increase. Absolute inhibitory concentration (absIC) was calculated as the corresponding point between the 0% and 100% assay controls. Fifty % and 80% inhibition were defined by the controls for all the samples on the same plate. For example, the absIC50 or absIC80 would be the point at which the curve matches inhibition equal to exactly 50% or 80% of the 100% assay control relative to the assay minimum.

## Data Availability

all data in the manuscript is available

## Acknowledgments

***Biobanco de Enfermedades Infecciosas Colección COVID19* working group:** Yesica Longueira, María L. Polo, Melina Salvatori, Sabrina Azzolina, Yanina Ghiglione, Horacio Salomon, María Florencia Quiroga, Gabriela Turk and Natalia Laufer.

Authors are thankful to Dr. Florian Krammer and Dr. Felix Rey for reagents and protocols for protein expression and purification, to Ignacio Sanchez and Alejandra Tortorici for protein purification protocols, to Matias Ostrowski and Paula Pérez for help in optimizing neutralization assay protocols, to Drs. Alicia Mistchenko, Belen Bouzas, Marisa Gimenez, Nadia Ahmed, Marcelo Rodriguez Fermepin, Romina Musante, Alejandra Margari, Juan Stupka, Maria Alejandra Morales, Ana Buchovsky, Maria Eugenia Bernardi and Mariela Aranda for recommendations during the COVIDAR validation process, and Dr. Laura Bover and the CPC19 group for help in protocol dissemination of antibody titrations. We are also grateful to Drs. Leandro Burgos Pratx and Ventura Simonovich for providing convalescent plasma samples and for discussion about COVID-19 convalescent plasma management, and to Drs. Jorge Geffner and Ana Fernandez-Sesma for insightful advice in COVIDAR applications. Thanks also go to Julieta Portillo, Marcos Olivera and Jorge Daniele from the Leloir Institute.

## Funding

The COVIDAR project was funded by the CONICET through the Fondo para la Convergencia Estructural del Mercosur (FOCEM) and Agencia Argentina de Promoción Científica y Tecnológica PICT2017-1717 Annex COVID-19 to AVG. Founds were also provided by Fundación Williams and Asociación Civil SAND for COVIDAR and Serokit production and distribution.

## Notes

### Competing Interest Statement

The authors have declared no competing interest.

### Author Declarations

Bioethics Committee of Fundacion Instituto Leloir (HHS IRB # 00007572) (CBFIL Protocol #35)

